# Towards routine long-read sequencing for rare disease: a national pilot study on chromosomal rearrangements

**DOI:** 10.1101/2023.12.15.23299892

**Authors:** Jesper Eisfeldt, Adam Ameur, Felix Lenner, Esmee Ten Berk de Boer, Marlene Ek, Josephine Wincent, Raquel Vaz, Jesper Ottosson, Tord Jonson, Sofie Ivarsson, Sofia Thunström, Alexandra Topa, Simon Stenberg, Anna Rohlin, Anna Sandestig, Margareta Nordling, Pia Palmebäck, Magnus Burstedt, Frida Nordin, Eva-Lena Stattin, Maria Sobol, Panagiotis Baliakas, Marie-Louise Bondeson, Ida Höijer, Kristine Bilgrav Saether, Lovisa Lovmar, Hans Ehrencrona, Malin Melin, Lars Feuk, Anna Lindstrand

**Affiliations:** Department of Molecular Medicine and Surgery and Center for Molecular Medicine, Karolinska Institutet, Stockholm, Sweden; Department of Clinical Genetics and Genomics, Karolinska University Hospital, Stockholm, Sweden; Science for Life Laboratory, Karolinska Institutet Science Park, Solna, Sweden; Department of Immunology, Genetics and Pathology, Uppsala University, Uppsala, Sweden; Science for Life Laboratory, Uppsala University, Uppsala, Sweden; Department of Clinical Genetics and Genomics, Sahlgrenska University Hospital, Gothenburg, Sweden; Division of Clinical Genetics, Department of Laboratory Medicine, Lund University, Lund, Sweden; Department of Clinical Genetics, Pathology and Molecular Diagnostics, Office for Medical Services, Region Skåne, Lund, Sweden; Department of Laboratory Medicine, Institute for Biomedicine, Sahlgrenska Academy, University of Gothenburg, Gothenburg, Sweden; Department of Clinical Genetics, University Hospital, Linköping, Sweden; Division of Cell and Neurobiology, Department of Biomedical and Clinical Sciences, Linköping University, Linköping, Sweden; Department of Medical Bioscience, Medical and Clinical Genetics, Umeå University, Umeå, Sweden; Department of Pharmacology and Clinical Neurosciences, Umeå University, Umeå, Sweden; Genomic Medicine Center Karolinska, Karolinska University Hospital, Stockholm, Sweden

**Keywords:** long-read genome sequencing, HiFi sequencing, de novo assembly, chromosomal rearrangements, structural variants, clinical diagnostics, rare diseases

## Abstract

**Background:** Clinical genetic laboratories often require comprehensive analysis of chromosomal rearrangements/structural variants (SVs) which can range from gross chromosomal events, such as translocations and inversions, to supernumerary ring/marker chromosomes, and small deletions or duplications. To fully understand the complexity of a specific event and its associated clinical consequences, it is imperative to locate the breakpoint junctions and to resolve the derivative chromosome structure. This task, however, often surpasses the capabilities of conventional short-read sequencing technologies. In contrast, emerging long-read sequencing techniques present a compelling alternative for clinical diagnostics.

**Methods:** Here, the Genomic Medicine Sweden Rare Diseases (GMS-RD) consortium explored the utility of HiFi Revio long-read whole genome sequencing (lrGS) for clinical digital karyotyping of SVs nationwide. The first 16 samples included in this study were collected from all health care regions in Sweden. We established a national pipeline and a shared variant database for variant calling and filtering. The included validation samples cover a spectrum of simple and complex SVs including inversions, translocations and copy number variants.

**Results:** The results from the lrGS analysis match the reported karyotype for 14/16 individuals and 12 known SVs were mapped at nucleotide resolution. A complex rearrangement on chromosome 15 was identified only through read depth analysis and two chromosome 21 rearrangements remained undetected, one of which was mosaic. The average read length ranged from 8.3-18.8 kb and the coverage was >20x for all samples. *De novo* assembly resulted in a limited number of contigs per individual (N50 range 6-86 Mb) clearly separating the two alleles in most cases, enabling direct characterization of the chromosomal rearrangements.

**Conclusions:** In a national pilot study, we successfully demonstrated the utility of HiFi Revio lrGS as a clinical analysis of chromosomal rearrangements. Based on our results we propose a five-year plan for the wider implementation of lrGS for rare disease diagnostics in Sweden.

## BACKGROUND

Although short-read (sr) genomic analysis approaches such as exome and genome sequencing (GS) have been highly successful in identifying disease-causing genetic variants for diagnostic purposes, the primary focus of analysis remains on single nucleotide variations (SNVs) and small insertions-deletions (INDELs) (1). In contrast, the calling and interpretation of structural chromosomal rearrangements is more challenging (2, 3). Such events, collectively called structural variants (SVs), are defined as genetic variants larger than 50 bp. When SVs involve a repositioning of genetic material either within a chromosome or between chromosomes, they are also referred to as chromosomal rearrangements. These encompass both recurrent and non-recurrent copy number variations (CNVs, deletions and duplications) and balanced chromosome abnormalities (translocations, inversions, and insertions). Furthermore, there is growing evidence that complex SVs, which contain multiple breakpoint junctions or consist of more than one simple structural variation in *cis*, are more prevalent than previously assumed (4). A thorough analysis of such events detailing the DNA breakpoints at the nucleotide level is crucial for comprehending the disease-causing and rearrangement-generating mechanisms. This level of understanding is necessary for effective personalized clinical management and genetic counselling (5).

While srGS shows great promise as a first-line diagnostic test, with potential to capture pathogenic SVs previously identified by traditional methods (6), it still has limitations. The existing srGS SV pipelines lack the ability to produce high-quality genome assemblies necessary to resolve complex disease-causing SVs. In this context, long-read genome sequencing (lrGS) has emerged as a promising alternative with potential to capture the complete scope of structural genomic variation in the human genome (7, 8). Indeed, lrGS provides a comprehensive approach for characterizing various types of SVs identified in a clinical genetic laboratory, whether discovered through traditional methods or srGS. With the introduction of lrGS using highly accurate consensus reads (HiFi reads), the quality of SNV and INDEL calls from lrGS data has increased dramatically and enables characterization of the full spectrum of genetic variation from SNVs to SVs in a single sequencing experiment. Due to the several kb long sequence reads, lrGS offers unique possibilities to study regions in the human genome that are not easily captured by other genomics technologies, such as highly repetitive or homologous regions. Furthermore, sequencing of the native DNA also provides methylation information. With decreasing cost, it is increasingly attractive to use lrGS in unsolved rare disease cases. Yet, lrGS differs markedly from short-read sequencing and various factors should be considered before its clinical implementation. These include the quality and quantity of DNA starting material, intricate specifics related to sequencing including read quality, total coverage, and read length, as well as the complexities tied to data analysis, visualization, and the eventual clinical interpretation.

Recognizing the promise and challenges of lrGS, we embarked on a national pilot study using the PacBio Revio system. We leveraged on Genomic Medicine Sweden (GMS), a national initiative designed to implement precision medicine throughout the regionally organized healthcare system (9) with the aim to validate lrGS for clinical digital karyotyping of unresolved SVs nationwide. The included samples originated from all university hospital regions in Sweden and the DNA was not necessarily collected using lrGS-optimized protocols. This approach mirrored the typical sample quality and preparation found in most of Sweden’s hospitals and biobanks today, thereby giving us a realistic view of the potential and challenges of lrGS in clinical diagnostics.

## METHODS

### Study subjects

All Swedish university regions are affiliated to the GMS working group for rare diseases (GMS-RD). In a nationwide lrGS pilot study samples from individuals with seemingly complex SVs were recruited from all healthcare regions. The multicenter study was approved by the Swedish Ethical Review Authority (2019–04746) and written informed consent was obtained from each participating individual or their respective legal guardians.

Altogether, 16 samples were sequenced including eleven proband singletons, one duo, and one trio. For two samples (P8.1 and P8.2) a high molecular preparation had been performed prior to lrGS using the Nanobind CBBX kit (Pacific Biosciences, Menlo Park, CA, USA) and following protocol Nanobind HMW DNA extraction – mammalian whole blood (PN 102-573-500, REV01), while the remaining DNA samples were retrieved from clinical biobanks and obtained using standard extraction protocols. Details on the included individuals are given in Table 1.

**Table 1:**
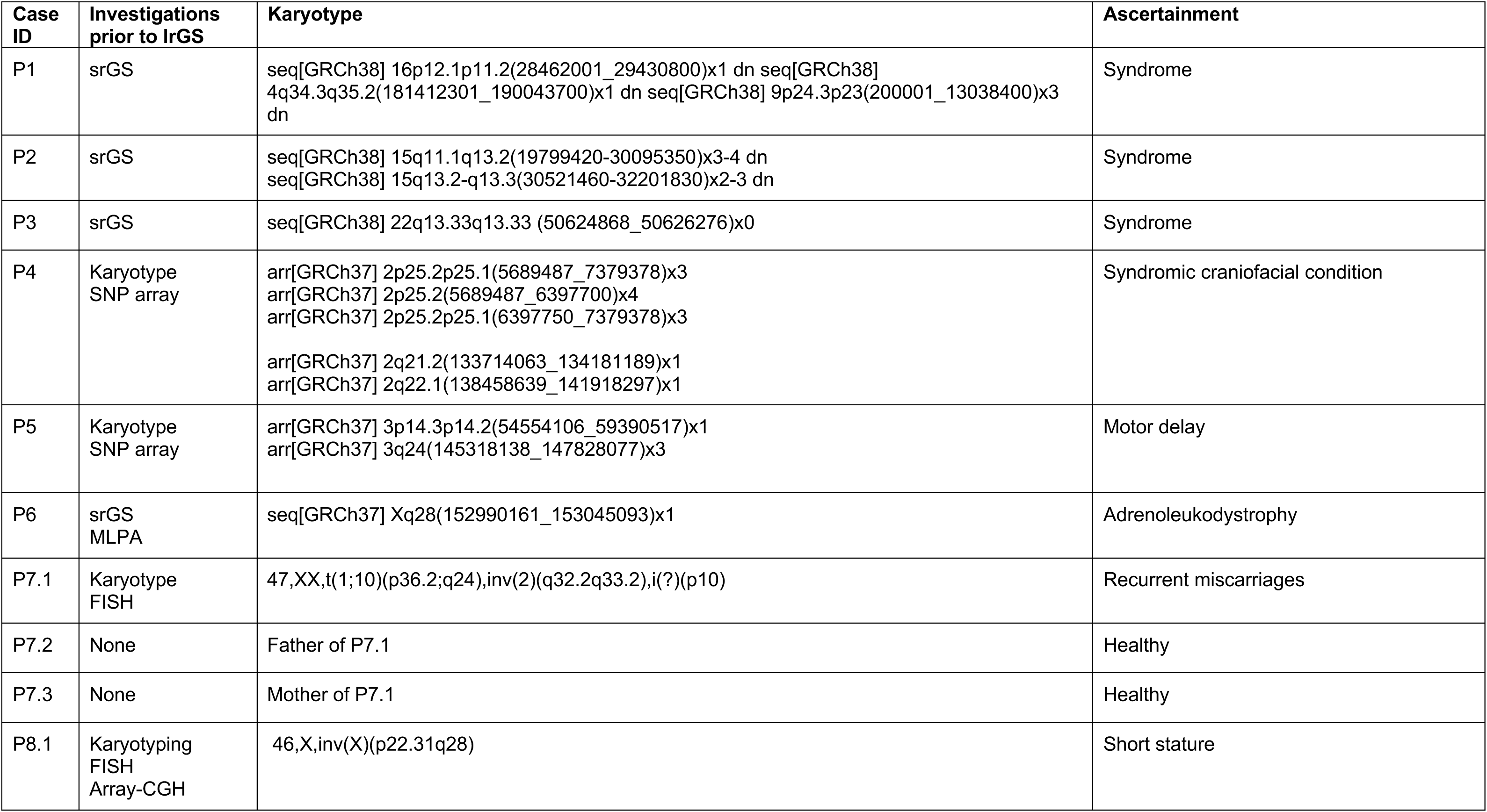
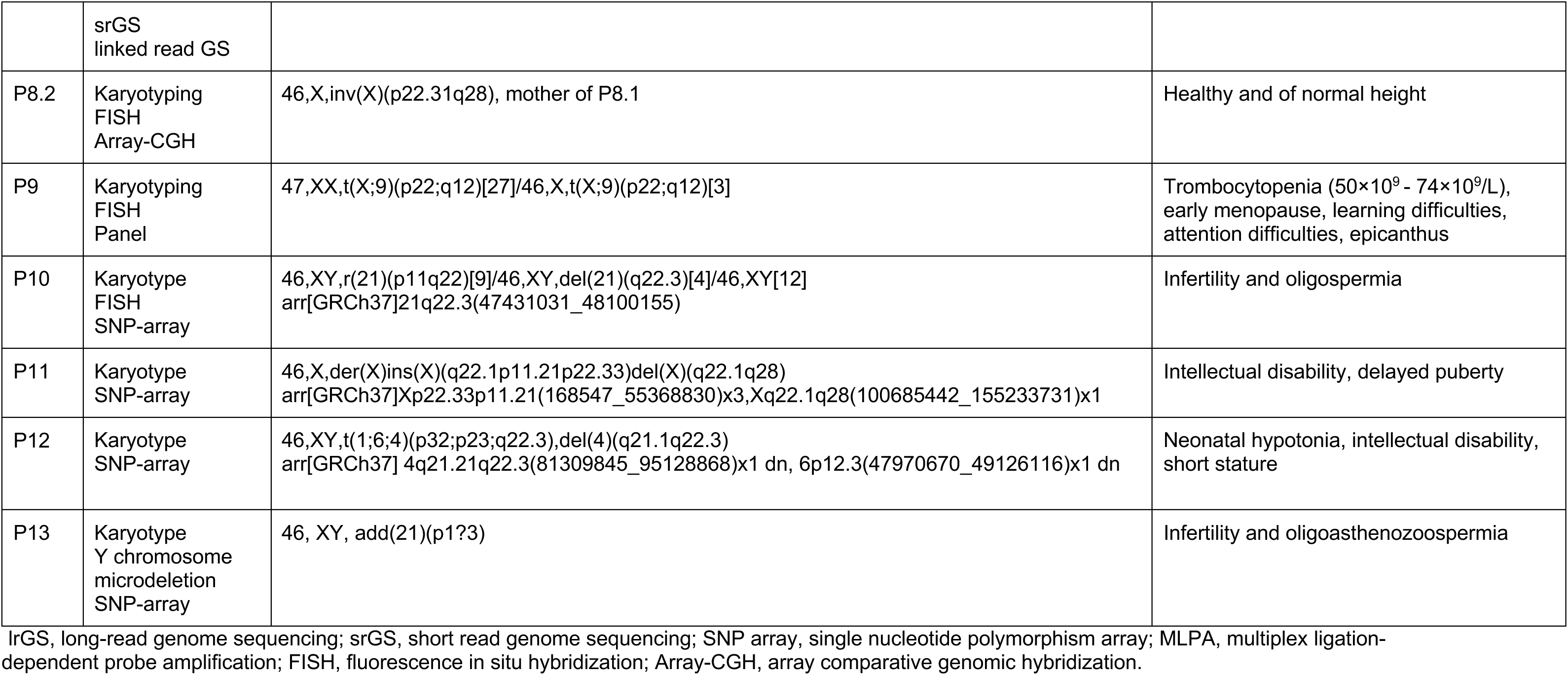
Karyotypes and mode of ascertainment of included cases.

### Long-read genome sequencing

The DNA samples were fragmented to 15-20 kb using Megaruptor 3 (Diagenode). PacBio SMRTbell library construction was performed using the SMRTbell Template prep kit 3.0. SMRTbell libraries were size-selected either using AMPure beads or by the gel-based systems pippinHT (Sage Science) or SageElf (Sage Science). The library preparation procedure is described in the protocol “Preparing whole genome and metagenome sequencing libraries using SMRTbell prep kit 3.0” from PacBio. The SMRTbell library sizes and profiles were evaluated using Fragment Analyzer (Agilent Technologies). PacBio sequencing was performed on the PacBio Revio system with 24 h movie time. Each SMRTbell library was sequenced on a 25M SMRT cell.

### Analysis of long-read genome data

The first 13 samples were run with skierfe (https://github.com/genomic-medicine-sweden/skierfe) commit d834d3c, and three individuals (P11-13) were run with skierfe commit bd11d1d. Additional analyses were then carried out on top of these results. Briefly, the PacBio HiFi reads were aligned to both GRCh38 and the T2T-CHM13v2.0 reference using minimap2 (version 2.26) (10) and SAMtools (version 1.17) (11). SNVs and INDELs were called with DeepVariant 1.5.0 (12) and phased using WhatsHap 1.7 (13). SVs were called using Sniffles (14), which was run according to previously published parameters (7), and CNVs were called using HiFiCNV (0.1.6b or 0.1.7) (https://github.com/PacificBiosciences/HiFiCNV). Quality metrics were gathered using fastqc 0.11.9 (http://www.bioinformatics.babraham.ac.uk/projects/fastqc/), cramino 0.9.7 (15) and mosdepth 0.3.3 (16). Phased assemblies were generated using hifiasm (version 0.19.5-r587) (17). The resulting *de novo* assemblies were aligned to hg38 and T2T-CHM13 reference using minimap2 (version 2.26) (10), SNVs were called using HTSBOX (version r345) (11) and counted using bcftools (version 1.17) (11), SVs were called using SVIM-asm using diploid mode for phased assemblies and haploid mode for non-phased assemblies (18) and quality control was performed using QUAST (version 5.0.2) (19).

### Identification and characterization of structural variants

Prior to SV characterization, the Sniffles SV calls were filtered based on size and allele frequencies. The size filtering was performed using bcftools (11), excluding all SV calls smaller than 2 kbp. The frequency filtering was performed using SVDB (20). First, a frequency database was constructed using the calls from all 16 individuals sequenced (Table 1). Next, we annotated the remaining calls using that database, and variants present in more than one unrelated individual were removed. Lastly, the remaining SV calls were inspected using IGV, and characterized as described previously (7).

## RESULTS

### High-quality long-read genome data is obtained from clinical DNA samples

All individuals were sequenced on one Revio 25M SMRTcell, generating at least 20X coverage of high-quality (HiFi) reads for all samples (range 19.9 - 35.5X), with mean read length ranging from 8.3-18.8 kb (Figure 1; Additional file 1: Table S1). Two samples (P8.1 and P8.2) were obtained from high-molecular weight DNA extraction of fresh blood, enabling for a more efficient size selection during library preparation. Gel-based size-selection was performed not only on those two samples but also on three regular DNA samples (P11, P12 and P13). Contrary to expectations, all five samples achived similar levels of high coverage and longer read lengths, with P13 reaching the highest overall throughput of >35 X coverage and the best assembly results. This indicates that the regular DNA extractions are suitable for HiFi sequencing and can be used to generate the best possible data from a single SMRTcell. Variant calling resulted in an average of 4.9 million single nucleotide variants (SNVs) and small INDELs per individual (Additional file 1: Table S2; Additional file 2: Document S2). This number of SNVs is similar to what is typically found by human srGS analysis. We further performed *de novo* assembly for all individuals and the contig N50 values are shown in Figure 1C.

**Figure 1:**
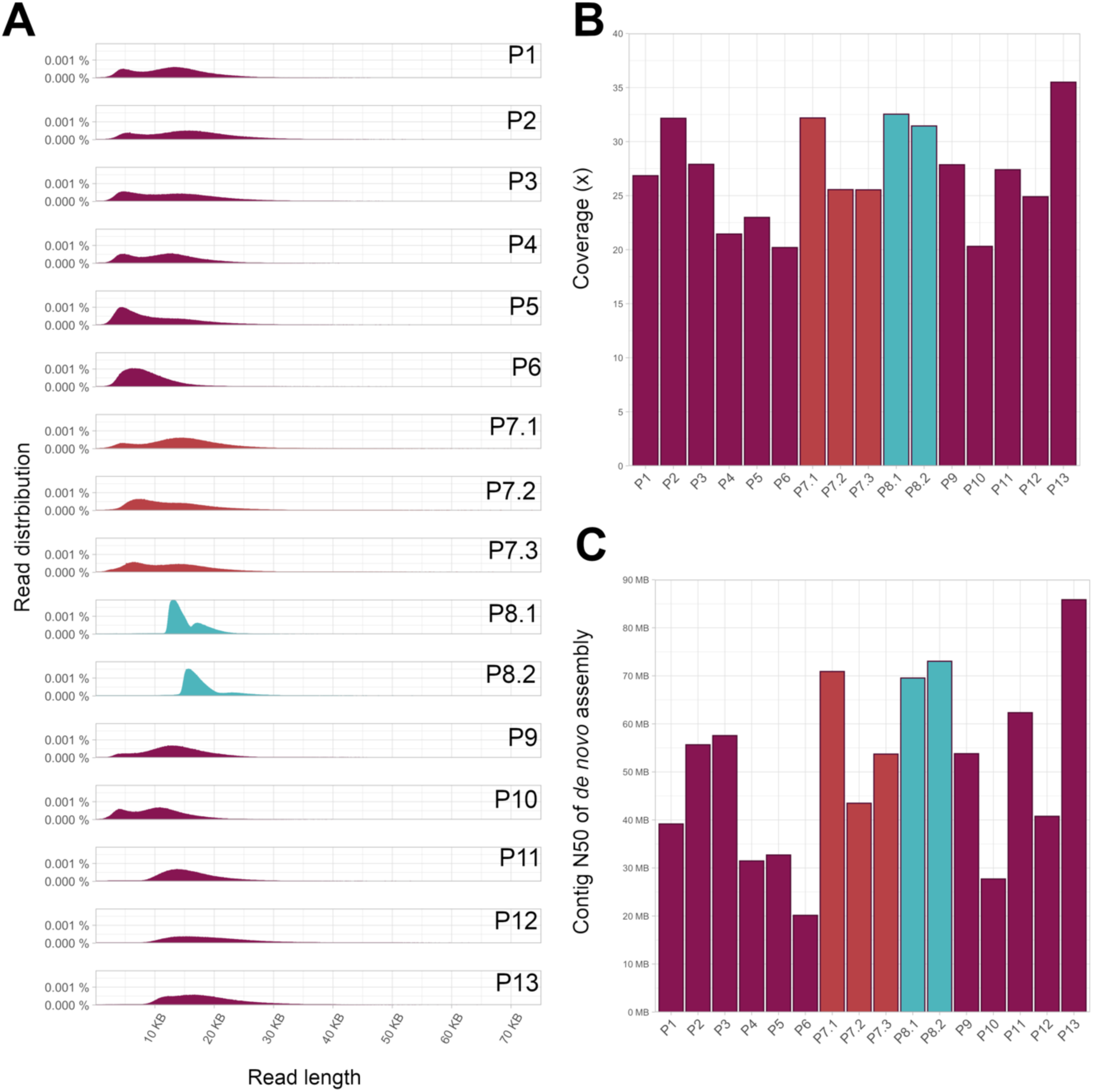
Quality measures of lrGS. Read length distribution (left). Coverage (top right) and N50 of *de novo* assembly (bottom right).

### Identification of chromosomal genomic rearrangements

The lrGS analysis could identify 11 of the 13 unique chromosomal rearrangements that were present in the 16 individuals sequenced. Of the two rearrangements that eluded detection, one was mosaic (P10) and the second affected the acrocentric p-arm of chromosome 21, a known repetitive genomic region (P13) (Table 1). The mosaic tri- and tetrasomy on chromosome 15 in individual P2 was characterized as a copy number gain (between 2 and 5) in the long-read CNV-calls, although a clear distinction between the tri- and tetrasomy could not be made. While the starting point could not be distinguished from the centromere, the location of the end point was approximated between chr15:32176000 – 32346000, with the exact breakpoint located within a segmental duplication (Additional file 1: Figure S1).

The remaining samples were then subdivided depending on whether they carried an inter-chromosomal or intra-chromosomal rearrangement. The HGVS nomenclature for the resolved events is given in Table 2.

**Table 2:** HGVS results of cases with identified BPJs subdivided into simple and complex rearrangements.

| Sample ID | HGVS karyotype | SV structure |
| --- | --- | --- |
| **Translocations and inversions** |  |  |
| P1 | chr4:g.qter_181414917delins[chr9:g.pter_13043905]<br>chr16:g.28442001_29508000del | t(4;9)<br>DEL |
| P7.1 P7.3 | chr1:g.19783172_qterdelins[chr10:g.pter_95395335]<br>chr10:g.pter_19783105delins[chr1:g.95395328_qter] | t(1;10) |
|  | chr2:g.189694535_202576085inv | inv(2) |
| P9 | chr9:g.pter_(75862011)delins[chrX:g.3044233_qter]<br>chrX:g.3044228_qterdelins[chr9:g.pter_(75862011)] | t(X;9) |
| P12 | chr4:g.80390384_qterdelins[chr6:g.pter_20052376inv]<br>chr1:g.58205788_qterdelins[chr4:g.94218568_106505089inv;chr6:20052374_qter]<br>chr1:g.pter_58205785delins[chr4:g.106505092_qterinv]<br>chr6:g.48002694_49160076 del | t(1;6;4)<br>DEL |
| **Complex rearrangements** |  |  |
| P3 | chr22:g.50618055_50626277delins[chr22:g.50624361_50624868inv] | DEL-INV-DEL |
| P4 | chr2:g.7246507_7734100delins[chr2:g.5620671_6248808inv;<br>chr2:g.5549092_7241289inv;chr2:g.5858296_6246303] | DUP-TRIP-QUAD-TRIP-DUP-NML-DEL |
|  | chr2:g.132954122_141163928delins[chr2:g.133434126_137703180inv] | DEL-INV-DEL |
| P5 | chr3:g.54518907_80981660delins[chr3:g.59407900_63614153inv;<br>chr3:g.135767535_135891958;chr3:g.63614153_80981660inv;<br>chr3:g.145600155_148110490] | DEL-INV-NML-DUP-NML-DUP |
| P6 | chrX:g.153724706_153724706delins[chr9:g.97596764_. inv] | DEL-INS-DEL |
| P8.1 | chrX:g.9768910_154113036delins[chrX:g.9420014_154208530inv] | DUP-INV-DUP |
| P8.2 | chrX:g.9768910_154113036delins[chrX:g.9420014_154208530inv] | DUP-INV-DUP |
| P11 | ChrX:g.101431832_qterdelins[chrX:g.pter_55349282<br>inv] | DUP-NML-DEL |

#### Translocations and inversions

Three translocations and one inversion were characterized in four individuals (Figure 2; Table 2) with a mother and daughter both carrying the same two balanced events (P7.1 and P7.3). All variants were fully resolved, but the translocation between chromosome X and 9 (P9) was only detectable using the Telomere-to-Telomere (T2T) assembly for analysis. A secondary analysis in hg38 revealed a call of a t(X;5) in that sample, indicating that the breakpoint 9 region is missing in GRCh38. In individual P1 the three structural variants (two deletions and one duplication) detected prior to lrGS were in fact two separate rearrangements; one recurrent 16p deletion and one unbalanced translocation between chromosome 4 and chromosome 9 (Figure 2). Finally, we were able to delineate a complex chromosome translocation involving chromosomes 1, 4 and 6 (P12). The analysis detected four breakpoint junctions (BPJs), a 13.8 MB deletion on chromosome 4 as well as an independent 1.2 MB deletion on chromosome 6 located centromeric of the chromosome 6 breakpoint (Figure 1D, Table 2, Table 3).

**Figure 2:**
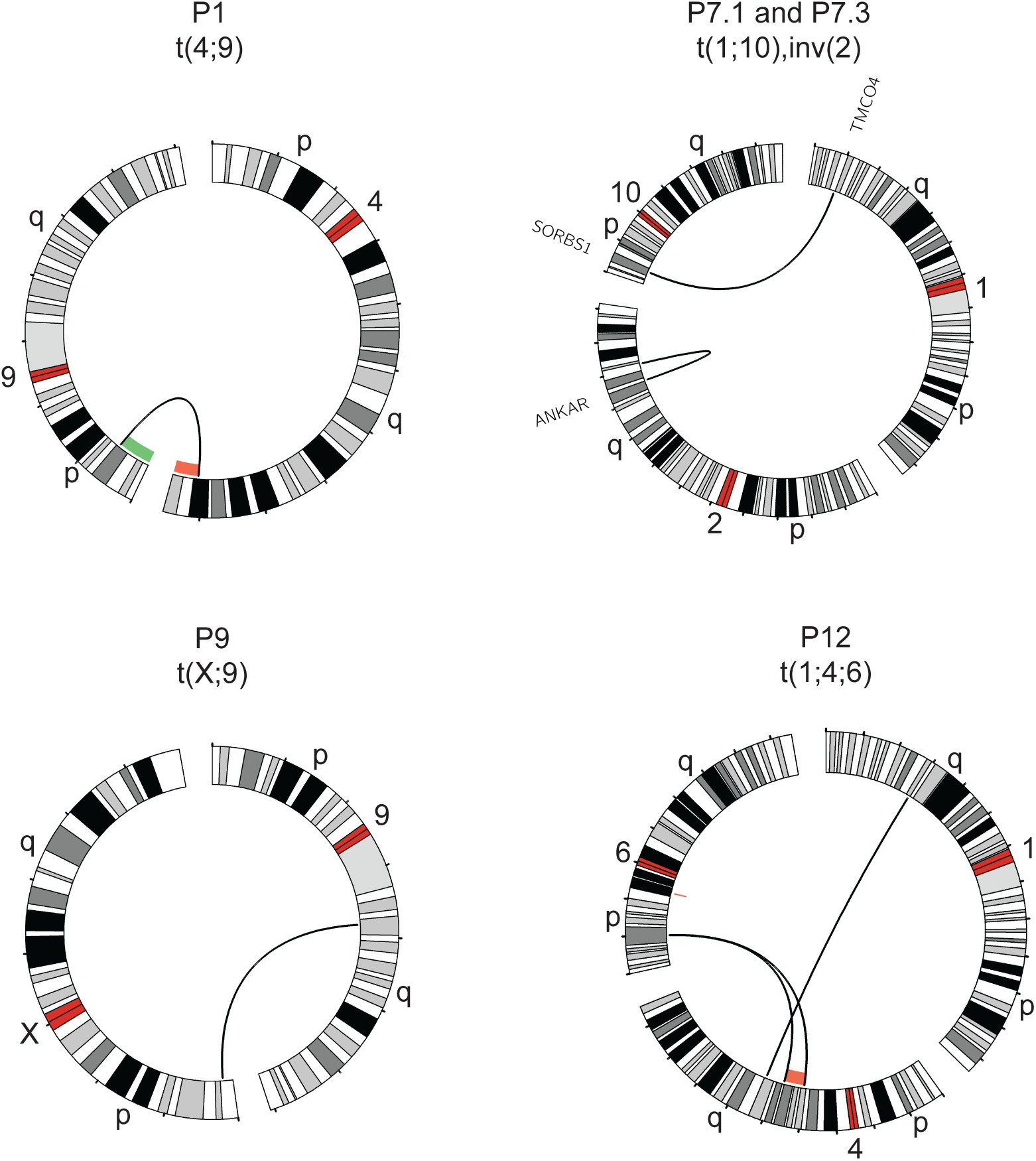
Translocations and inversions identified with lrGS. Circos plots of rearrangements detected in four cases using lrGS: a t(4;9) in P1, a t(1;10) and inv(2) (P7.1 and P7.3), a t(X;9) (P9) and a t(1;4;6) (P12). A green/red line indicates copy number gain/loss, respectively. Genes disrupted are indicated at the breakpoint.

**Table 3:** Junction characteristics of identified rearrangements, one case was only detected in T2T.

| Sample ID | BPJs | ChrA | posA | chrB | posB | Strand | Gene posA | Gene posB | Breakpoint insertions | Repeat A | Repeat B | Microhomology |
| --- | --- | --- | --- | --- | --- | --- | --- | --- | --- | --- | --- | --- |
| **Hg38** |  |  |  |  |  |  |  |  |  |  |  |  |
| P1 | 1 | 4 | 181414917 | 9 | 13043905 | +/- | - | - | - | L1 | L1 | - |
|  | 1 | 16 | 28455071 | 16 | 29476612 | +/+ | - | - | - | *AluJo* | *SVA_D* | - |
| P3 | 2 | 22 | 50618055 | 22 | 50624868 | +/- | - | **ARSA** | 2nt | tigger1 | MIR | - |
|  |  | 22 | 50624361 | 22 | 50626277 | +/- | **ARSA** | **ARSA** | - | *Alusp* | - | 2nt |
| P4 | 4 | 2 | 5549092 | 2 | 5858296 | +/- | *XR_001739261.1* | - | - | - | L1PA7 | 4nt |
|  |  | 2 | 5620671 | 2 | 7241289 | +/+ | - | - | 9nt | - | - | - |
|  |  | 2 | 6246303 | 2 | 7734100 | +/+ | - | - | - | L1M5 | - | 2nt |
|  |  | 2 | 6248808 | 2 | 7246507 | +/- | - | - | - | L1ME4b | - | 3nt |
|  | 2 | 2 | 132954122 | 2 | 137703180 | +/- | *NCKAP5* | - | - | MLT1K | L1PA16 | 1nt |
|  |  | 2 | 133434126 | 2 | 141163928 | +/- | *NCKAP5* | *LRP1B* | - | - | - | 11nt |
| P5 | 5 | 3 | 54518907 | 3 | 63614153 | +/- | *CACNA2D3* | *SYNPR* | - | - | MamRep38 | - |
|  |  | 3 | 59407900 | 3 | 135767535 | +/- | *XR_002959675.1* | - | - | - | MER101 | 1nt |
|  |  | 3 | 63614153 | 3 | 145600155 | +/- | *SYNPR* | - | - | MamRep38 | L2A | - |
|  |  | 3 | 80981660 | 3 | 135891958 | +/- | - | - | - | MSTA-int | L2a | 1nt |
|  |  | 3 | 80981660 | 3 | 148110490 | +/+ | - | - | 1nt | MSTA-int | MER58A | - |
| P6 | 2 | 9 | 97596764 | X | 153779639 | +/- | *TMOD1* | - | - | - | - | 1nt |
|  |  | 9 | 97598236 | X | 153724706 | +/- | *TMOD1* | *BCAP31* | 12nt | *AluSX1* | - | - |
| P7.1<br>P7.3 | 4 | 1 | 19783172 | 10 | 95395335 | +/- | *TMCO4* | *SORBS*<br>*1* | 7nt | MIRb | - | - |
|  |  | 1 | 19783105 | 10 | 95395328 | +/- | *TMCO4* | *SORBS*<br>*1* | 5nt | MIRb | - | - |
|  |  | 2 | 189694535 | 2 | 202576085 | +/- | - | *ANKAR* | 1nt | - | L1PA17 | - |
|  |  | 2 | 189694533 | 2 | 202576083 | +/- | - | *ANKAR* | - | - | L1PA17 | 1nt |
| P8.1<br>P8.2 | 2 | X | 9420014 | X | 154113037 | +/- | XR_0017<br>55796.1 | - | 28nt | *AluJr* | *AluSx1* | - |
|  |  | X | 9768909 | X | 154208530 | +/- | GPR143 | - | 31nt | *AluSz6* | *AluJo* | - |
| P11 | 1 | X | 55349282 | X | 101431831 | +/- | - | *ARMCX*<br>*4* | - | *AluY* | *AluY* | 290nt |
| P12 | 5 | 4 | 80390384 | 6 | 20052376 | +/- | CFAP299 | - | - | *L1MA9* | *AluSq2* | 1nt |
|  |  | 4 | 94218568 | 6 | 20052374 | +/- | *SMARCA*<br>*D1* | - | - | *AluSz* | *AluSq2* | 1nt |
|  |  | 1 | 58205787 | 4 | 106505089 | +/- | *DAB1* | - | - | - | *THE1D* | - |
|  |  | 1 | 58205785 | 4 | 106505092 | +/- | *DAB1* | - | - | - | *THE1D* | 2nt |
|  |  | 6 | 48002694 | 6 | 49160076 | +/+ | *PTCHD4* | - | - | *L1PA4* | - | 3nt |
| T2T |  |  |  |  |  |  |  |  |  |  |  |  |
| P9 | 2 | 9 | 75862011* | X | 3044233 | +/- | - | - | - | (AATGG)n | ERVl-MaLR | 2nt |
|  |  | 9 | 75862011* | X | 3044228 | +/- | - | - | 990 nt | (TTCCA)n | ERVL-MaLR | - |

BPJ, breakpoint junction; chr, chromosome; pos, position; nt, nucleotide; Bold text indicates a disease-causing gene. \*=estimated location

#### Complex intrachromosomal rearrangements

In addition to the complex translocation discussed above, six intrachromosomal complex rearrangements were detected and resolved in five unrelated individuals, all of which were unbalanced (Table 2). Notably, the same inv(X) was detailed in a mother and daughter pair (P8.1 and P8.2). The highest complexity level was identified in P4 where two complex events were identified, one DUP-TRIP-QUAD-TRIP-DUP-DEL and one DEL-INV-DEL (Figure 3A; Table 2). The complexity found in P4 was tightly followed by P5, who carries a complex rearrangement consisting of two duplications, one deletion, as well as an inverted copy number neutral segment (Figure 3B; Table 2). In contrast, the simplest rearrangements consisted of DEL-INV-DEL (P3 and P4) or DEL-INS-DEL (P6) (Additional file 1: Figure S2). Finally, we resolved a terminal invDUP-NML-DEL on chromosome X (P11), mediated by matching *Alu*Y elements and harboring a 291 long stretch of microhomology (Table 3).

**Figure 3:**
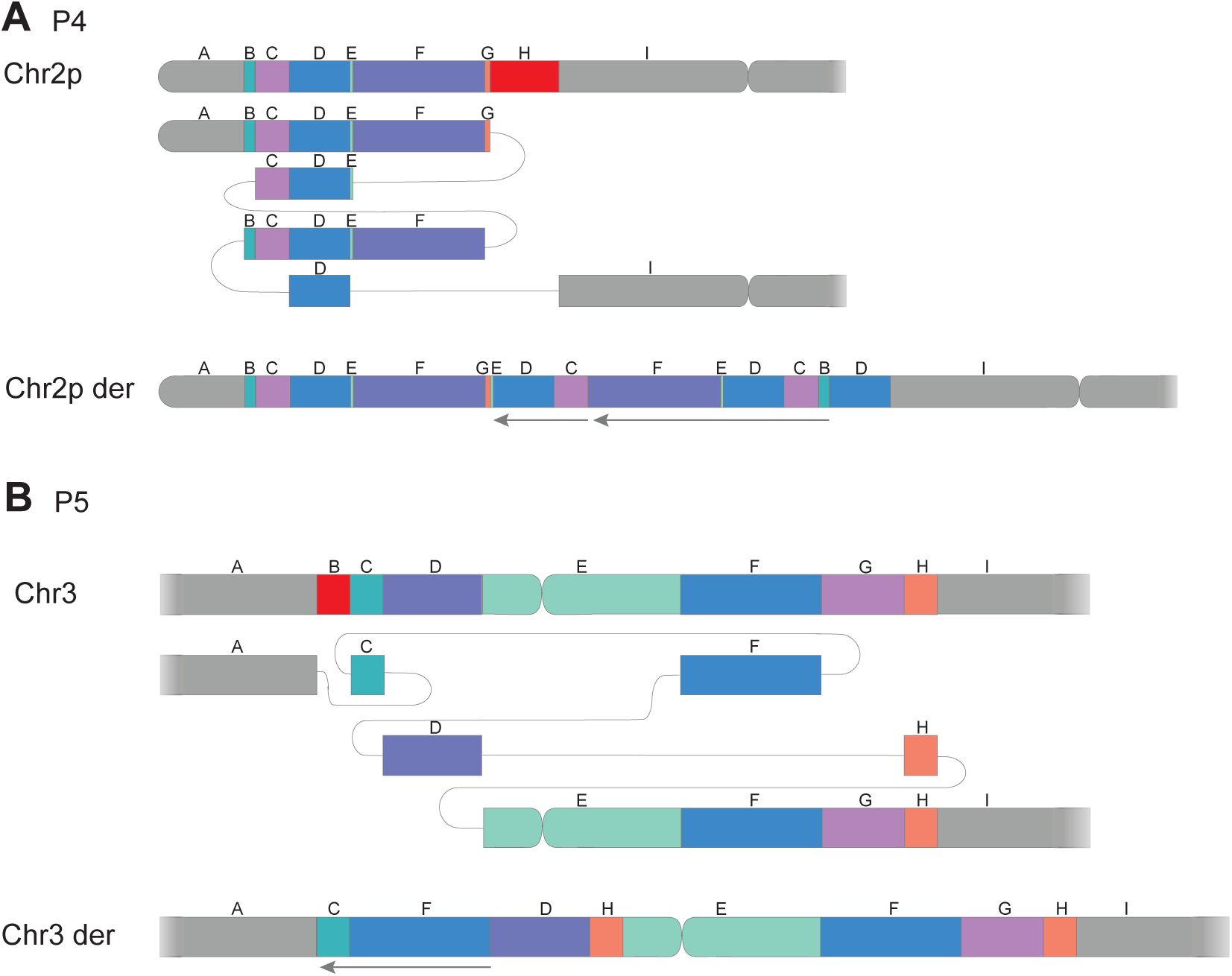
Subway plots of two complex rearrangements: A) A complex dup-trip-quad-trip-dup-del on chromosome 2 observed in P4. B) A clustered CNV on chromosome 3 detected in P5. Deleted segments shown in red, arrows mark inverted segments.

### Breakpoint junction analysis

In eleven of the rearrangements, the BPJs were characterized at nucleotide resolution, allowing for a detailed analysis of insertions, repeat elements and microhomology and disrupted genes (Table 3; Additional file 1: Table S3). Comparing the Sniffles SV calls to the calls from our *de novo* assembly pipeline, we find that the *de novo* workflow detects 30 of the 31 BPJs detected by Sniffles (Table 2, Table 3; Additional file 1: Table S3). Notably, we find a great diversity of such genetic signatures in the analyzed cases. In three rearrangements, the breakpoints contain matched repeat elements: L1 in the t(4;9) (P1) and various *Alu* elements in the inv(X) (P8.1 and P8.2) as well as in P11. In individual P4 (Table 3) microhomology of 1-11 nucleotides (nt) was observed in five of the six BPJs and the final BPJ contains a nine nt insertion. Insertions larger than 2nt were detected in three additional rearrangements (Table 3); a complex translocational insertion (P6) (Additional File 1: Figure S2), a t(1;10) (P7.1 and P7.3) (Figure 2) and a t(X;9) (P9) (Figure 2). The final two rearrangements, a DEL-INV-DEL on chr22 (P3) (Additional File 1: Figure S2) and a complex DEL-INV-NML-DUP-NML-DUP on chr3 (P5, Figure 3B), showed blunt ends that contained neither microhomologies, matched repeats, or larger insertions (Table 3).

### Characterization of background structural variants

Next, we assessed the SV burden in all the 16 samples of the GMS-RD lrGS cohort. On average, we detected 23 814 SVs per individual that were subdivided into complex (0.05%), deletions (41%), duplications (1%), insertions (51%), inversions (1%) and translocations (5%) (Figure 4A). In general, the detected SVs were small, with 82% of SV below 1000bp and a peak is observed at 300bp representing the *Alu* insertions and deletions (Figure 4B). Most variants are present in more than one of the 13 analyzed individuals and only 30% are unique (Figure 4C). Finally, when comparing to the SweGen SV database (21) containing background SVs in 1000 Swedish individuals assessed from srGS data, approximately 70% of the lrGS SV calls are novel and not present in the SweGen dataset (Figure 4D).

**Figure 4:**
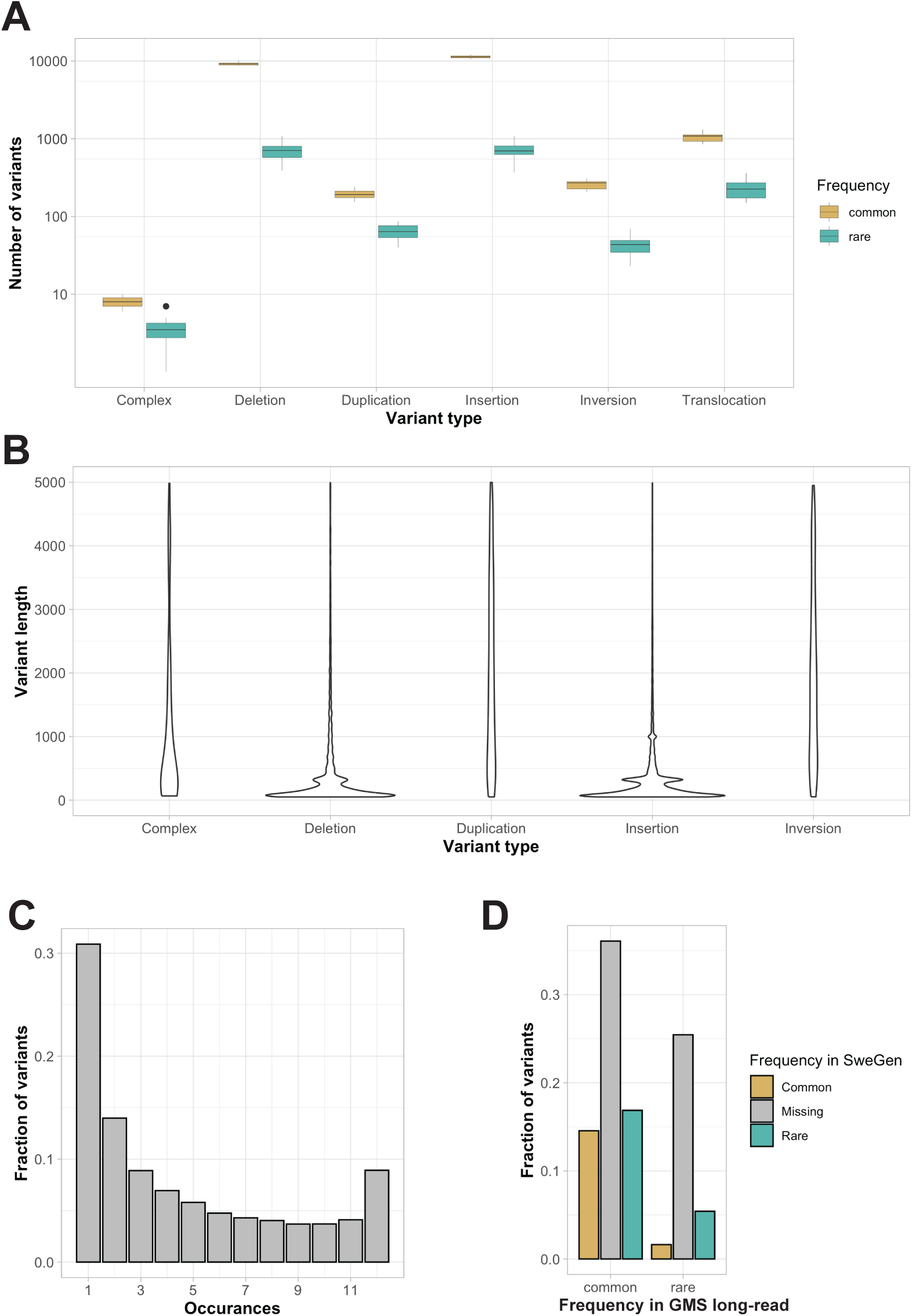
Characterization of background SVs. A) Boxplot illustrating the number of SVs per type (common in yellow and rare in green). B) Violin plot of SV length per SV type (excluding SV > 5kbp). C) Allele frequency histogram. D) Comparison of allele frequencies between the GMS long-read cohort and SweGen srGS SV database (common in yellow, missing in gray and rare in green).

## DISCUSSION

In this national collaborative project, we not only demonstrated the usefulness of lrGS as a clinical follow up analysis of SVs detected by other methods, but we have also taken the first steps towards clinical long-read diagnostics of rare diseases in our nation. By utilizing the expertise in our different regions, we have developed a comprehensive workflow covering sample collection, DNA preparation as well as sequencing and data analysis.

We successfully resolved 12 unique chromosomal rearrangements, but in three instances, lrGS failed to detect the BPJs. Specifically, lrGS missed two chromosome 21 rearrangements (P10, P13) and one case of mosaic partial tri- and tetrasomy on chromosome 15 (P2), which was only revealed through read depth analysis. This underscores the necessity of including read depth callers for a comprehensive lrGS analysis. The coverage in the two undetected events (20X and 35X respectively for P10 and P13) might have affected our sensitivity. It is difficult to predict what coverage would have been required since that would depend on the degree of mosaicism, type of structural variant as well as genomic context. It is likely that at least 40-60X would have been needed in these cases.

By resolving the structure of the BPJs in 12 rearrangements (Table 4) we uncovered additional complexities compared to the original analysis important for a complete clinical interpretation. In addition, we identified diverse mechanisms of SV formation. Matched repeats were present at the BPJs in three rearrangements (P1, P8, P11), a pattern indicative of non-allelic homologous recombination (NAHR) or *Alu*/*Alu*-mediated rearrangements (P8, P11) (22–24). In five complex SVs from four individuals (P3, P4, P6 and P7) both microhomology and insertions were observed at the breakpoints (Table 4), features indicative of replicative mechanisms such as Fork Stalling and Template Switching (FoSTeS)/ microhomology-mediated break-induced replication (MMBIR) (25, 26). However, the largest insertion, 990 nt present at the BPJ of the t(X;9) in P9, consists mainly of TTCCA repeats that may be due to background variation in this repeat, and not caused through the formation of the translocation.

In a broader context, the current case series showcases how SVs represent a promising start for the introduction of lrGS into clinical diagnostics. However, a more complete transition to lrGS will take some time. As such, GMS-RD have drafted a five-year roadmap, during which new lrGS diagnostic tests successively will be implemented in Sweden (Figure 5). To accomplish this, there will be a need to streamline the workflows for DNA extraction, library preparation, sequencing, bioinformatic analysis and variant interpretation. Based on this pilot study, we here highlight some of the main challenges and potential solutions for the wider implementation of lrGS into clinical routine.

**Figure 5:**
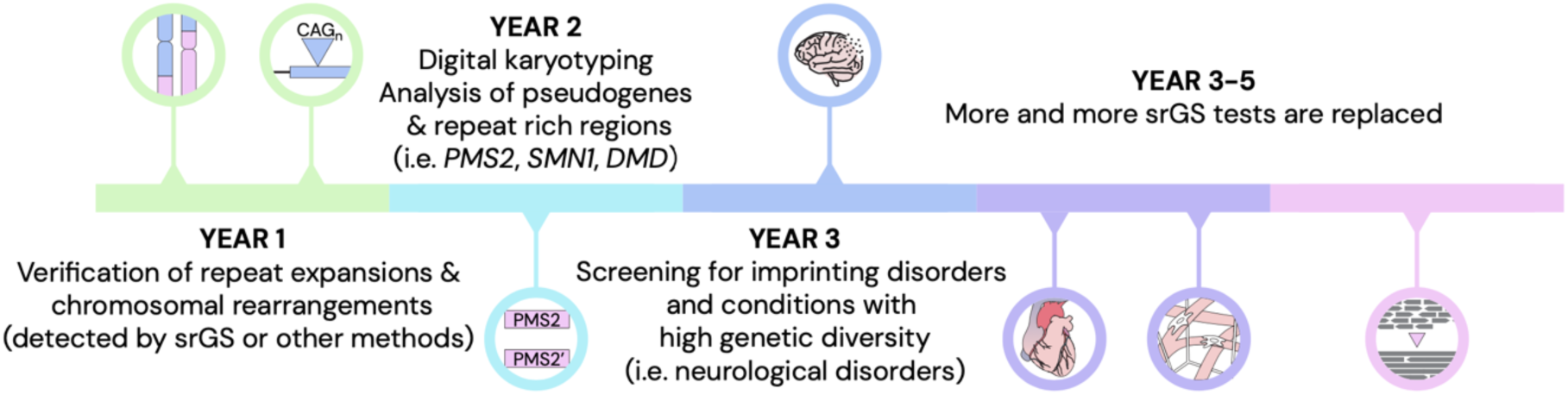
Towards long-read genome sequencing as a first tier diagnostic test in rare disease. A five-year timeline with expected development of long-read sequencing in the clinical setting.

Starting with DNA extraction, our results show that lrGS with PacBio Revio performs well with blood samples extracted in routine diagnostics. We obtained >20x coverage for all the samples, which is likely enough for calling of both small and large variants (27, 28). The two samples where HMW-DNA was extracted from fresh blood (P8.1, P8.2) gave slightly more data on average, but we also obtained high yields from the three routine DNA samples (P11, P12, P13) where we had enough DNA to perform gel-based size selection (Additional file 2: Document S2). For routine clinical implementation, we therefore propose to use the regular DNA extraction protocols that are already implemented at the hospitals, which will reduce the turn-around time and cost.

The second step is library preparation which was performed manually. Although feasible for a small number of samples, it will not scale in clinical routine, where 1000’s of rare disease patients and relatives are expected to be tested yearly in Sweden (9). We therefore face the need to implement automated library preparation on liquid handling systems. This will be challenging for certain library preparation steps, such as the shearing and size selection that are not part of a typical srGS library preparation.

Regarding the long-read sequencing the main limitations are capacity and costs. The PacBio Revio system used in this study has a theoretical capacity of around 1300 human genomes per year which is ∼15 times less than the Novaseq X platform. Nevertheless, advancements in technology may lower costs and increase throughput, allowing for more patient testing. Introducing barcoding could also enable sequencing multiple samples per run, improving efficiency. Particularly, for certain rare disease patient groups, such as those with neurological disorders caused by repeat expansions, targeted gene panels might be a more cost-effective approach (29, 30).

Long-read sequencing generates large amounts of data that need to be processed with streamlined bioinformatics pipelines. For clinical implementation, we need robust and efficient pipelines. Moreover, since bioinformatics is a rapidly evolving research area, it is crucial that the pipelines can be modified as new software emerges. We see great value in processing samples from different hospitals with a similar analysis pipeline, since this will enable joint downstream analyses and comparisons between larger patient groups. For this purpose, we have established a common pipeline (https://github.com/genomic-medicine-sweden/skierfe) for the lrGS analysis of rare disease patients in Sweden. All regions in the country have access to the pipeline and can contribute to its continuous development.

Finally, the called variants undergo prioritization and interpretation. In this pilot the focus was on SVs and for all cases there was prior information about the chromosomal rearrangement. In the future, when lrGS is applied as a first line test for rare disease diagnostics the data analysis will be more challenging. In particular, there is a need to establish a national lrGS reference dataset for effective filtering of non-pathogenic variants. To this end, we are planning to sequence a cross-section of the Swedish population. The initial reference dataset will consist of at least 100 individuals capturing most of the common variants in the population. Ideally, ethnic minority groups should also be included although it might be possible to obtain such data through international collaborations. In addition to a national reference dataset, other lrGS resources such as HPRC (31) and ONT 1000G (32) will be valuable. It may also be necessary to utilize multiple reference genomes, or even a human pangenome graphs(31), since we noticed here that T2T-CHM13 (33) enables detection of SV breakpoints not seen in GRCh38.

Variant calling is followed by variant prioritization, enabling identification of the pathogenic variant(s). During this process, visualization is an important tool. IGV (34), often used for short-reads, is mainly designed for the visualization of small variants (INDELs and SNVs), and for SVs identified by lrGS alternative methods will be required. In this study, we generated subway and circos plots for this purpose, but these might not be suitable for all types of variants. Particularly, specialized tools should be implemented to visualize tandem repeats (35, 36).

When a new candidate variant has been detected and visually inspected, its role in the disease needs to be established. We expect that many of the novel variants detected through lrGS will be of unknown significance, since they have not been observed with previous genome technologies, and therefore it will be important to obtain good annotations and functional predictions. To some extent, the same tools as for srGS can be used for functional prediction, but it may be a challenge to understand potential consequences of complex SVs, especially those located within non-coding regions. For that reason, it may be necessary to focus on the most obvious results and leave some of the uncertain diagnoses for later. As the databases and annotations grow, we can then revisit those patients and hopefully give a correct diagnosis.

## Conclusions

Here we demonstrate that by coordinating our local efforts and working together in the GMS Rare Diseases consortium we were able to build the tools and workflows necessary to validate lrGS for digital karyotyping in the entire nation. Even though there is still quite some work to be done for full clinical utility of lrGS, our preliminary results show that there is no question that lrGS will provide benefit for rare disease diagnostics. With the plan we have laid out we hope to have the methods set up and running at scale for rare disease within a five-year period.

## Abbreviations

Array-CGH: array comparative genomic hybridization
BPJ: breakpoint junction
chr: chromosome
CNV: copy number variant
FISH: fluorescence in situ hybridization
FoSTeS: fork-stalling and template switching
GMS: Genomic Medicine Sweden
HMW: high molecular weight
lrGS: long-read genome sequencing
MLPA: multiplex ligation-dependent probe amplification
nt: nucleotide
pos: position
SNV: single nucleotide variant
srGS: short-read genomic sequencing
SV: structural variant
T2T: Telomere-to-Telomere.

## Declarations

## Data Availability

The PacBio HiFi data and individual variant calls will be available through the European '1+ Million Genomes' Initiative that is being set up. As a clinical case data will be stored at the National Genomics Platform managed by Genomic Medicine Sweden, accession number GMS-RD_00002. Data will be made available upon reasonable request through the corresponding authors.

## Acknowledgments

We are very grateful to the participating families. We would also like to thank UPPMAX for the use of computer infrastructure resources (project sens2022541) and the support from the National Genomics Infrastructure (NGI) Uppsala and Clinical Genomics Uppsala. Several authors of this publication are members of the European Reference Network on Rare Congenital Malformations and Rare Intellectual Disability ERN-ITHACA [EU Framework Partnership Agreement ID: 3HP-HP-FPA ERN-01-2016/739516]. We also acknowledge the Uppsala ALF office.

## Availability of data and materials

Analysis results supporting the conclusions of this article are included within the article and its additional files. The PacBio HiFi data and individual variant calls will be available through the European ’1+ Million Genomes’ Initiative that is being set up. As a clinical case data will be stored at the National Genomics Platform managed by Genomic Medicine Sweden, accession number GMS-RD_00002. Data will be made available upon reasonable request through the corresponding authors.

The following public databases and open-source software were used:

Genome Reference Consortium Human Build 38 (https://www.ncbi.nlm.nih.gov/datasets/genome/GCF_000001405.26/)

The Telomere-to-Telomere consortium CHM13 project T2T-CHM13v2.0 ( https://github.com/marbl/CHM13)

Assemblatron (https://github.com/J35P312/Assemblatron) BCFtools (https://samtools.github.io/bcftools/bcftools.html) (11)

Cramino (https://github.com/wdecoster/cramino) (15)

Deepvariant (https://github.com/google/deepvariant) (12)

FastQC (https://www.bioinformatics.babraham.ac.uk/projects/fastqc/)

Hifiasm (https://github.com/chhylp123/hifiasm) (17)

Hificnv (https://github.com/PacificBiosciences/HiFiCNV)

HTSBOX (https://github.com/J35P312/htsbox) (11)

Minimap2 (https://github.com/lh3/minimap2) (10)

Samtools (https://www.htslib.org/) (11)

Sniffles (https://github.com/fritzsedlazeck/Sniffles) (14)

SVDB (https://github.com/J35P312/SVDB) (20)

Mosdepth (https://github.com/brentp/mosdepth)

MultiQC (https://multiqc.info/)

Whatshap (https://whatshap.readthedocs.io/en/latest/) (13)

The SweGen dataset (https://swefreq.nbis.se/dataset/SweGen) (21)

## Authors’ contributions

JE, AA, MM, LL, HE, LF and AL conceptualized the study. JE, AA, FL, ETB, RV, LF and AL were major contributors to writing the manuscript. ME and IH performed extraction and QC analysis of DNA. JW, JO, TJ, SI, ST, AT, SS, AR, AS, MN, PP, MB, FN, ELS, MS, PB, MLB, LL, HE and AL provided patient samples and clinical information of patients. JE, AA, FL, ETB, KBS and LF performed bioinformatics analysis and development. All authors have read, edited and approved the final manuscript.

## Ethics approval and consent to participate

The Ethical Review Board in Sweden approved the study (ethics permit number 2019-04746). Written consent to participate was provided by the subject or their legal guardians. The research conformed to the principles of the Helsinki Declaration.

## Consent for publication

Written informed consent was obtained to publish.

## Supporting information

Additional file 1: Document S1 (Bioinformatic supplementary methods), Table S1 (Basic quality metrics of the samples included in this study), Table S2 (De novo assembly quality metrics), Table S3 (De novo assembly variant calling), Figure S1 (Read coverage across chromosome 15), Figure S2 (Subway plots of characterized rearrangements).

Additional file 2: Document S2 (MultiQC report)

