## Additional file 1 for "Towards routine long-read sequencing for rare disease: a national pilot study on chromosomal rearrangements"

**Supplementary Tables**

**Table S1:** Basic quality metrics of the samples included in this study

| **Sample ID** | **Mean coverage** | **Mean read length (bp)** | **Sequencing Yield (Gb)** |
|---|---|---|---|
| P1 | 26.5 | 12,307 | 82 |
| P2 | 31.7 | 14,896 | 98 |
| P3 | 27.5 | 12,563 | 85 |
| P4 | 21.2 | 11,233 | 66 |
| P5 | 22.7 | 9,567 | 70 |
| P6 | 19.9 | 8,343 | 62 |
| P7.1 | 31.7 | 14,265 | 98 |
| P7.2 | 25.2 | 12,277 | 78 |
| P7.3 | 25.2 | 12,284 | 78 |
| P8.1 | 32.1 | 15,104 | 99 |
| P8.2 | 31.0 | 17,612 | 96 |
| P9 | 27.5 | 13,285 | 86 |
| P10 | 20.1 | 9,849 | 63 |
| P11 | 27.4 | 15,716 | 90 |
| P12 | 24.9 | 18,810 | 84 |
| P13 | 35.5 | 17,940 | 119 |

Bp, base pairs; Gb, giga bases

**Table S2:** De novo assembly quality metrics

| Assembly | Contigs (n) | Total length (bp) | N50 (bp) | Genome Fraction (%) | Small variants(n) | SVs (n) | Indels |
|---|---|---|---|---|---|---|---|
| P1 hap1 | 952 | 3011730018 | 27656118 | 97.294 | 3573451 | 34337 | 706786 |
| P1 hap2 | 809 | 3020289601 | 27627387 | 97.003 | 3579905 | | 703549 |
| P1 no hap | 497 | 3055351291 | 39172056 | 97.545 | 3602598 | 33737 | 707649 |
| P2 hap1 | 528 | 3049271102 | 53162323 | 97.431 | 3625230 | 35135 | 713293 |
| P2 hap2 | 607 | 3022576100 | 30089741 | 97.345 | 3639562 | | 716270 |
| P2 no hap | 373 | 3077093992 | 55656714 | 97.596 | 3665279 | 34006 | 718203 |
| P3 hap1 | 805 | 2968923625 | 38003410 | 95.147 | 3629122 | 35288 | 714993 |
| P3 hap2 | 683 | 2955832984 | 26901484 | 94.768 | 3608950 | | 709099 |
| P3 no hap | 470 | 3070113002 | 57555714 | 97.867 | 3727047 | 32916 | 732983 |
| P4 hap1 | 2167 | 3009507143 | 10032076 | 96.786 | 3543724 | 33,371 | 723511 |
| P4 hap2 | 1870 | 2994360817 | 10443380 | 96.707 | 3600158 | | 729278 |
| P4 no hap | 990 | 3061337924 | 31471136 | 97.581 | 3649743 | 33158 | 734887 |
| P5 hap1 | 1410 | 3021926484 | 16791079 | 97.001 | 3564965 | 33505 | 710273 |
| P5 hap2 | 1442 | 3002452954 | 10513426 | 96.988 | 3559703 | | 710897 |
| P5 no hap | 780 | 3050115512 | 32705197 | 97.460 | 3621814 | 33780 | 717110 |
| P6 hap1 | 2891 | 2990264780 | 5665656 | 96.073 | 3589314 | 33140 | 734226 |
| P6 hap2 | 2786 | 2894053352 | 5660041 | 93.124 | 3516636 | | 718105 |
| P6 no hap | 1092 | 3059225159 | 20150227 | 97.822 | 3667936 | 32845 | 746927 |
| P7.1 hap1 | 543 | 3038562420 | 44240849 | 97.376 | 3620569 | 35664 | 708884 |
| P7.1 hap2 | 516 | 3037726858 | 49152777 | 97.415 | 3627346 | | 709222 |
| P7.1 no hap | 387 | 3070887463 | 70906275 | 97.538 | 3650945 | 34901 | 710859 |
| P7.2 hap1 | 1098 | 3004917728 | 26806592 | 96.139 | 3581699 | 34708 | 714973 |
| P7.2 hap2 | 909 | 2994455258 | 23607483 | 95.712 | 3581148 | | 715250 |
| P7.2 no hap | 579 | 3083744197 | 43485889 | 98.143 | 3649458 | 34144 | 727696 |

| | | | | | | | |
|---|---|---|---|---|---|---|---|
| P7.3 hap1 | 919 | 3007113736 | 29658224 | 97.315 | 3602776 | 35187 | 716780 |
| P7.3 hap2 | 785 | 3029744114 | 24927346 | 97.265 | 3628930 | | 716232 |
| P7.3 no hap | 514 | 3055007195 | 53717007 | 97.582 | 3660593 | 34144 | 720613 |
| P8.1 hap1 | 541 | 3024919811 | 56515313 | 97.460 | 3627237 | 35517 | 708068 |
| P8.1 hap2 | 571 | 3025639302 | 38174944 | 97.279 | 3594413 | | 706405 |
| P8.1 no hap | 367 | 3065271514 | 69545151 | 97.632 | 3643325 | 36161 | 709464 |
| P8.2 hap1 | 628 | 3027302868 | 62105149 | 97.384 | 3621918 | | 706580 |
| P8.2 hap2 | 526 | 3015816193 | 49998385 | 97.365 | 2469965 | | 491224 |
| P8.2 no hap | 415 | 3058931416 | 73038069 | 97.708 | 3667377 | 33791 | 711103 |
| P9 hap1 | 655 | 3034915555 | 54793259 | 97.408 | 3600299 | 33117 | 711583 |
| P9 hap2 | 706 | 3025770706 | 28512459 | 97.274 | 3570230 | | 2861175 |
| P9 no hap | 554 | 3066133149 | 53797994 | 97.617 | 3604444 | 33028 | 712078 |
| P10 hap1 | 2742 | 3014608061 | 8875707 | 96.653 | 3548895 | 35094 | 722959 |
| P10 hap2 | 2623 | 2886636947 | 7067025 | 92.984 | 3473909 | | 704156 |
| P10 no hap | 1301 | 3062985831 | 27709462 | 97.976 | 3617165 | 29860 | 732080 |
| P11 hap1 | 940 | 3033114741 | 40056718 | 96.571 | 3621609 | 34036 | 711084 |
| P11 hap2 | 884 | 2978655919 | 31312892 | 95.787 | 3525896 | | 699382 |
| P11 no hap | 597 | 3051400356 | 62335257 | 97.275 | 3647060 | 33749 | 714031 |
| P12 hap1 | 1300 | 3036918378 | 23705991 | 96.653 | 3610216 | 34670 | 750021 |
| P12 hap2 | 1047 | 2916967371 | 21936164 | 93.432 | 3550325 | | 730708 |
| P12 no hap | 746 | 3088070636 | 40766497 | 97.750 | 3699510 | 32626 | 759519 |
| P13 hap1 | 305 | 3021367031 | 58169784 | 96.398 | 3602439 | 34989 | 707153 |
| P13 hap2 | 278 | 2936874730 | 60498075 | 93.783 | 3563910 | | 694026 |
| P13 no hap | 276 | 3101975158 | 85852130 | 97.896 | 3677600 | 36840 | 716890 |

**Table S3:** De novo assembly variant calling

| Sample ID | ChrA | posA | chrB | posB | Found? |
|---|---|---|---|---|---|
| P1 | 4 | 181414917 | 9 | 13043905 | Yes |
| | 16 | 28454138 | 16 | 29 482 543 | Yes |
| P2 | - | - | - | - | - |
| P3 | 22 | 50618055 | 22 | 50624868 | Yes |
| | 22 | 50624361 | 22 | 50626277 | Yes |
| P4 | 2 | 5549092 | 2 | 5858296 | Yes |
| | 2 | 5620671 | 2 | 7241289 | No |
| | 2 | 6246303 | 2 | 7734100 | Yes |
| | 2 | 6248808 | 2 | 7246507 | Yes |
| | 2 | 132954122 | 2 | 137703180 | Yes |
| | 2 | 133434126 | 2 | 141163928 | Yes |
| P5 | 3 | 54518907 | 3 | 63614153 | Yes |
| | 3 | 59407900 | 3 | 135767535 | Yes |
| | 3 | 63614153 | 3 | 145600155 | Yes |
| | 3 | 80981660 | 3 | 135891958 | Yes |
| | 3 | 80981660 | 3 | 148110490 | Yes |
| P6 | 9 | 97596764 | X | 153779639 | Yes |
| | 9 | 97598236 | X | 153724706 | |

| | | | | | |
|---|---|---|---|---|---|
| P7.1 | 1 | 19783172 | 10 | 95395335 | Yes |
| | 1 | 19783105 | 10 | 95395328 | Yes |
| | 2 | 189694535 | 2 | 202576085 | Yes |
| | 2 | 189694533 | 2 | 202576083 | Yes |
| P7.2 | - | - | - | - | - |
| P7.3 | 1 | 19783172 | 10 | 95395335 | Yes |
| | 1 | 19783105 | 10 | 95395328 | Yes |
| | 2 | 189694535 | 2 | 202576085 | Yes |
| | 2 | 189694533 | 2 | 202576083 | Yes |
| P8.1 | X | 9420014 | X | 154113037 | Yes |
| | X | 9768909 | X | 154208530 | No |
| P8.2 | X | 9420014 | X | 154113037 | Yes, non-phased |
| | X | 9768909 | X | 154208530 | Yes, non-phased |
| P9 | 9 | 75862011* | X | 3044233 | Yes |
| | 9 | 75862011* | X | 3044228 | Yes |
| P10 | - | - | - | - | - |
| P11 | X | 101431832 | X | 55349282 | Yes |
| P12 | 4 | 80390384 | 6 | 20052376 | Yes |
| | 1 | 58205788 | 4 | 106505089 | Yes |

| | | | | | |
|---|---|---|---|---|---|
| | 6 | 20052374 | 4 | 94218568 | Yes |
| | 4 | 106505092 | 1 | 58205785 | Yes |
| | 6 | 48002694 | 6 | 49160076 | Yes |
| P13 | - | - | - | - | - |

**Supplementary Figures**

**Figure S1: Read coverage variation across chromosome 15.** Average read coverage per 1 MB segments of chromosome 15 for sample P2.

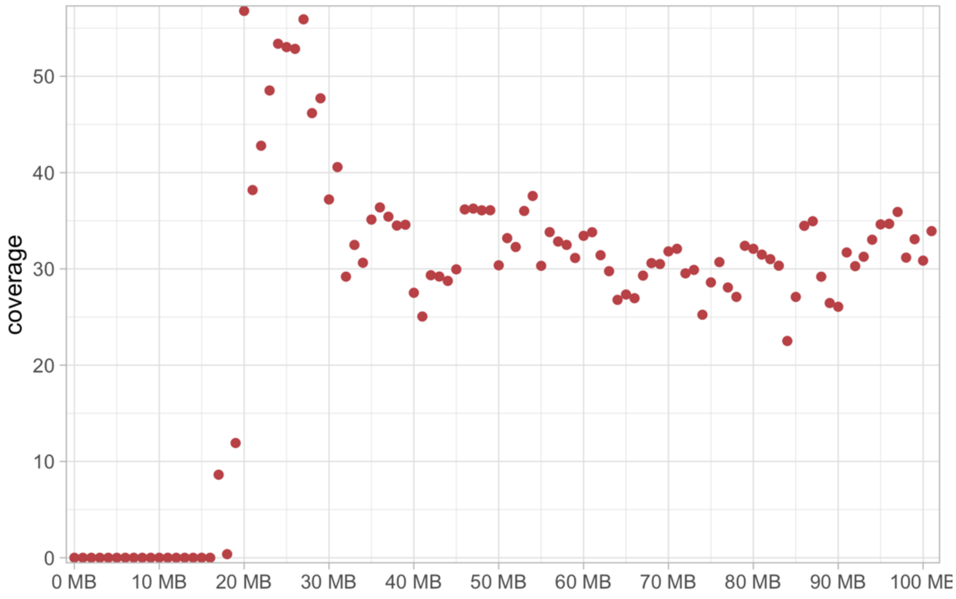

**Figure S2: Subway plots of characterized rearrangements.** Schematic illustrations of the resolved rearrangements in P3, P6, P4, P11 and P8. Red and pink segments are deleted in the patients and arrows show the inverted segments. Der: derivative chromosome.

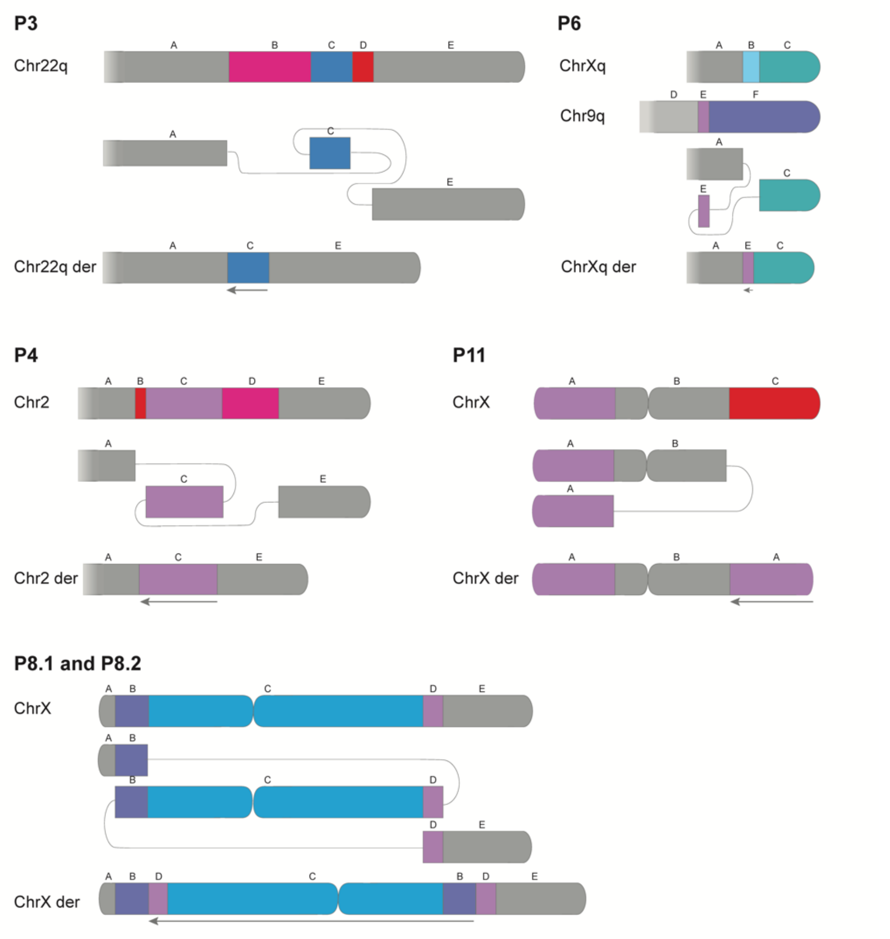


### SUPPLEMENTARY REFRENCES
